# Genetic association study of Preterm birth and Gestational age in a population-based case-control study in Peru

**DOI:** 10.1101/2023.11.22.23298891

**Authors:** Diana L. Juvinao-Quintero, Sixto E. Sanchez, Tsegaselassie Workalemahu, Nelida Pinto, Liming Liang, Michelle A. Williams, Bizu Gelaye

## Abstract

Preterm birth (PTB) is an adverse pregnancy outcome affecting ∼15 million pregnancies worldwide. Genetic studies have identified several candidate loci for PTB, but results remain inconclusive and limited to European populations. Thus, we conducted a genome-wide association study (GWAS) of PTB and gestational age at delivery (GA) among 2,212 Peruvian women. PTB cases delivered ≥ 20 weeks’ but < 37 weeks’ gestation, while controls delivered at term (≥ 37 weeks but < 42 weeks). After imputation (TOPMED) and quality control, we assessed the association of ∼6 million SNPs with PTB and GA using multivariable regression models adjusted for maternal age and the first two genetic principal components. *In silico* functional analysis (FUMA-GWAS) was conducted among top signals detected with an arbitrary *P* < 1.0×10^-5^ in each GWAS. We sought to replicate genetic associations with PTB and GA identified in Europeans, and we developed a genetic risk score for GA based on European markers. Mean GA was 30 ± 4 weeks in PTB cases (N=933) and 39 ± 1 in the controls (N=1,279). PTB cases were slightly older and had higher C-sections and vaginal bleeding than controls. No association was identified at genome-wide level. Top suggestive (*P* < 1.0×10^-5^) signals were seen at rs13151645 (*LINC01182*) for PTB, and at rs72824565 (*CTNNA2*) for GA. Top PTB variants were enriched for biological pathways associated with polyketide, progesterone, steroid hormones, and glycosyl metabolism. Top GA variants were enriched in intronic regions and cancer pathways, and these genes were upregulated in the brain and subcutaneous adipose tissue. In combination with non-genetic risk factors, top SNPs explained 14% and 15% of the phenotypic variance of PTB and GA in our sample, but these results need to be interpreted with caution. Variants in *WNT4* associated with GA in Europeans were replicated in our study. The genetic risk score based in European markers, was associated with a 2-day longer GA (R^2^=0.003, *P*=0.002) per standard deviation increase in the score in our sample. This genetic association study identified various signals suggestively associated with PTB and GA in a non-European population; they were linked to relevant biological pathways related to the metabolism of progesterone, prostanoid, and steroid hormones, and genes associated with GA were significantly upregulated in relevant tissues for the pathophysiology of PTB based on the *in-silico* functional analysis. None of these top variants overlapped with signals previously identified for PTB or GA in Europeans.

## INTRODUCTION

Preterm birth (PTB)—the premature onset of labor before 37 weeks of gestation [1–3]—affects 10% (∼ 15 million) of newborns globally [1, 4] with the majority of them (80%) occurring in the Sub-Saharan and South Asia region [1]. PTB is one of the leading causes of neonatal morbidity and mortality, and the leading cause of infant mortality before 5 years [5]. Preterm infants who survive are at higher risk of short-term and long-term disability; they are more prone to suffer from respiratory diseases [6, 7], necrotizing enterocolitis [8], neurodevelopmental impairment [9,

10], hypertension [11, 12], and glucose intolerance [13]. The pathogenesis of PTB remains largely unknown but is recognized as a multifactorial syndrome [14], influenced by multiple inflammatory-driven processes like microbial-induced inflammation, decidual hemorrhage, vascular disease, and disruption of the maternal-fetal immune tolerance, among others [2, 14]. Common PTB risk factors include maternal sociodemographic (ethnicity, low socioeconomic status, education, marital status, working conditions, and age) and behavioral characteristics (psychosocial distress, depression, substance abuse, smoking) [2, 15]; pregnancy history (previous PTB, preeclampsia, gestational diabetes), present pregnancy status (multigravida, intrauterine infection, uteroplacental ischemia or hemorrhage, uterine overdistention, cervical length, other comorbidities), and biological factors (nutritional status, BMI, inflammatory markers, fetal fibronectin) and genetic markers [2].

Twin studies have demonstrated the genetic contribution to PTB, with a reported heritability of ∼27% to 36% relative to maternal genetic effects [15], while evidence from genome-wide association studies (GWAS) have revealed several single nucleotide polymorphisms or SNPs with small effects on PTB and gestational duration [15–17]. Some of the challenges in studying the genetics of PTB are their reliance on both, the maternal and fetal genetic effects [18], the heterogeneity in the clinical definition of PTB, measurement error in the assessment of gestational age [18], and overall differences in the genetic risk observed between populations [15]. These challenges have limited the possibility of replicating signals across studies. GWAS in European ancestry individuals have identified six loci associated with gestational duration (*EBF1*, *EEFSEC*, *AGTR2*, *WNT4*, *ADCY5*, and *RAP2C*), four of which were also associated with PTB (*EBF1*, *EEFSEC*, and *AGTR2*) based on maternal genetic effects. These loci were linked to relevant biological pathways of PTB, including uterine development, maternal nutrition, and vascular control [16]. Using the fetal genotype, one variant in the *SLIT2* gene was associated with PTB in a Finnish population [19]. The *SLIT2* gene and its receptor *ROBO1* were upregulated in the placenta of PTB infants and participated in the regulation of genes involved in inflammation, decidualization, and fetal growth [19]. A recent GWAS using the maternal genotype from UK samples identified several SNPs and gene transcripts associated with spontaneous PTB, like the SNP rs14675645 (*ASTN1*), and transcripts from the microRNA-142 and *PPARG1-FOXP3* gene associated with PPROM [20]. Overall, these markers were linked to inflammation and immune response pathways [20]. In a trans-ethnic GWAS using the fetal genotype [15], two intergenic SNPs were identified in association with extreme PTB cases (< 30 weeks of gestation) in populations from African (chr 1, rs17591250) and American (chr 8, rs1979081) ancestries, which differed from SNPs previously identified in Europeans, and could not be replicated in additional cohorts [15]. Lastly, GWAS investigating gene-environment interactions in the risk of PTB have identified SNPs in *PTPRD* and *COL24A1* where the genotype interacts with maternal lifetime stress and pre-pregnancy BMI, respectively, in increasing the risk of spontaneous PTB in African Americans [21, 22].

So far, genetic evidence points towards a population-specific genetic risk for PTB and gestational age [15, 17, 18], which may be reflective of the interaction between the genotype and specific environmental risk factors. However, most genetic studies have focused on European ancestry populations, with a limited representation of admixture populations like those in the Americas [15], limiting our ability to identify the mechanisms associated with PTB in minority populations at increased risk. Thus, we conducted the present study to identify maternal genetic variants associated with PTB and gestational age at delivery (GA) in a case-control study from Lima, Peru. We hypothesize that unique markers associated with PTB and GA will be identified, reflecting distinct mechanistic pathways to PTB in this admixture population.

## RESULTS

### Population Characteristics

As shown in Table 1 women were, on average, 28 years of age, with most of them being multiparous, and had normal pre-pregnancy BMI, and low educational attainment. Only a small proportion of these mothers reported smoking tobacco or using illicit drugs during pregnancy (< 2%), while 17% of them reported using alcohol. Compared to controls, PTB cases were older, more likely to deliver through C-section, and had poorer self-reported health in pregnancy. PTB cases were also more likely to be preeclamptic, report vaginal bleeding during pregnancy, and deliver infants with lower birthweight compared to control mothers. Additional charactristics of the study participants are provided in S1 Table.

**Table 1.** Sociodemographic characteristics of study participants in Lima, Peru (N= 2,212).

|  | Overall<br>Sample<br>(N= 2,212)<br>n (%) | Preterm-Births<br>(N= 933)<br>n (%) | Full-term births<br>(N= 1,279)<br>n (%) | P* |
| --- | --- | --- | --- | --- |
| Maternal age (years), Mean (SD) | 28 (7) | 29 (7) | 28 (7) | 0.01 |
| Maternal age by groups |  |  |  |  |
| 18-19 | 211 (10) | 74 (8) | 137 (11) | 0.03 |
| 20-29 | 1,095 (50) | 462 (50) | 633 (50) |  |
| 30-34 | 444 (20) | 181 (20) | 263 (21) |  |
| ≥ 35 | 438 (20) | 207 (22) | 231 (18) |  |
| Gestational age (weeks), Mean (SD) | 35 (5) | 30 (4) | 39 (1) | <0.001 |
| PPROM, yes (%) | 515 (24) | 379 (42) | 136 (11) | <0.001 |
| Primiparous, yes (%) | 680 (31) | 273 (29) | 407 (32) | 0.20 |
| Mode of delivery |  |  |  |  |
| Natural | 822 (37) | 287 (31) | 535 (42) | <0.001 |
| C-section | 1385 (63) | 644 (69) | 741 (58) |  |
| Maternal Education |  |  |  |  |
| ≤ high school | 1,557 (71) | 634 (69) | 923 (73) | 0.036 |
| > high school | 636 (29) | 290 (31) | 346 (27) |  |
| Marital status |  |  |  |  |
| Married or living with partner | 1,896 (86) | 786 (85) | 1,110 (87) | 0.11 |
| Not married or living alone | 304 (14) | 141 (15) | 163 (13) |  |
| Employed during pregnancy |  |  |  |  |
| No | 999 (45) | 414 (45) | 585 (46) | 0.60 |
| Yes | 1,198 (55) | 510 (55) | 688 (54) |  |
| Pre-pregnancy BMI (kg/m<sup>2</sup>), Mean (SD) | 25 (5) | 25 (5) | 25 (5) | 0.04 |
| Pre-pregnancy BMI (categories) |  |  |  |  |
| Underweight (<18.5) | 53 (2.5) | 29 (3.2) | 24 (1.9) | 0.12 |
| Normal weight (18.5–24.9) | 1,196 (56) | 512 (57) | 684 (55) |  |
| Overweight (25 – 29.9) | 614 (29) | 241 (27) | 373 (30) |  |
| Obese (≥ 30) | 275 (13) | 118 (13) | 157 (13) |  |
| Planned pregnancy, yes (%) | 733 (33) | 291 (31) | 442 (35) | 0.08 |
| Health perception, Fair/poor (%) | 1,230 (56) | 603 (65) | 627 (50) | <0.001 |
| Alcohol use, yes (%) | 370 (17) | 168 (18) | 202 (16) | 0.20 |
| Tobacco smoking, yes (%) | 25 (1) | 12 (1) | 13 (1) | 0.60 |
| Drug abuse, yes (%) | 4 (0.2) | 0 (0) | 4 (0.3) | 0.14 |
| Preeclampsia, yes (%) | 180 (8.2) | 90 (9.7) | 90 (7.1) | 0.03 |
| Vaginal bleeding, yes (%) | 432 (20) | 273 (29) | 159 (12) | <0.001 |
| Spontaneous Abortions, yes (%) | 616 (28) | 277 (30) | 339 (27) | 0.10 |
| Infant birthweight (g), Mean (SD) | 2,664 (1,169) | 1,656 (670) | 3,340 (867) | <0.001 |
| Child sex |  |  |  |  |
| Female | 993 (46) | 397 (43) | 596 (47) | 0.08 |
| Male | 1,189 (54) | 519 (57) | 670 (53) |  |

SD: standard deviation. PROM, Preterm premature rupture of membranes. \*Wilcoxon rank sum test; Pearson's Chi-squared test; Fisher's exact test.

### Ancestry Assessment

The PC analysis using combined genetic data from PAGE and the 1000 Genomes allowed us to determine the ancestry of PAGE participants, who generally clustered close to PEL samples (Peruvians from Lima, Peru), and a few others were dispersed among samples from other American regions (CLM Colombians, MXL Mexican American) (Fig 1). Using cleaned, directly genotyped genetic data from PAGE alone, we computed genetic PCs to use as covariates in genetic association analyses. We extracted the first 2 PCs explaining 56.5% and 9.65% of the total variation in the sample. Some dispersion was observed in the distribution of PAGE samples across these two genetic PCs.

**Fig 1.**
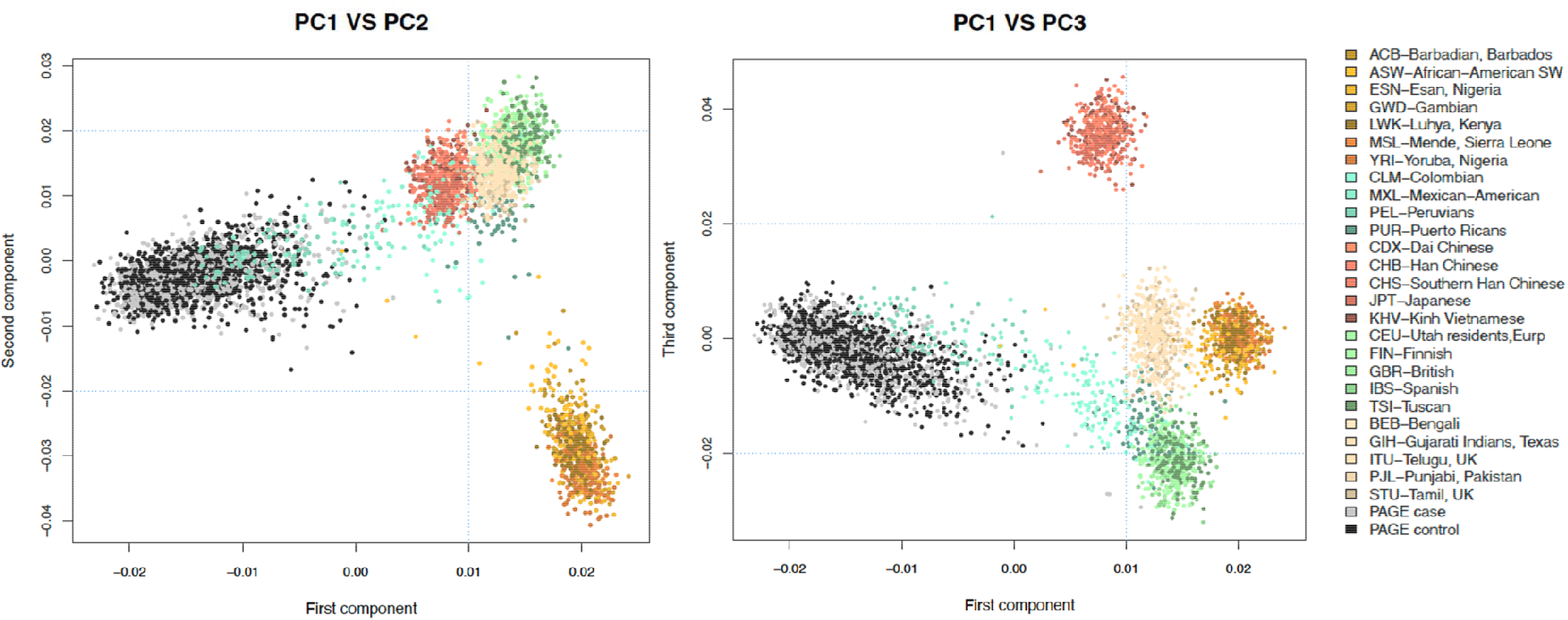
Genetic ancestry analysis showing the clustering of PAGE samples relative to global populations from different ancestries included in the 1000 Genomes project. More genetically related populations clustered more closely with each other than genetically distant populations. Most of the PAGE participants (black and grey dots) aligned close to the 1000 Genomes PEL samples (Peruvians from Lima, Peru), and few others were dispersed among samples from other populations in the Americas (CLM Colombians, MXL Mexican Americans). PC1-PC3: first three principal components. Subpopulations: ACB African Caribbean in Barbados, ASW African Ancestry in Southwest US, BEB Bengali in Bangladesh, CDX Chinese Dai in Xishuangbanna, CEU Utah residents with Northern and Western European ancestry, CHB Han Chinese in Beijing, CHS Southern Han Chinese, CLM Colombian in Medellin, ESN Esan in Nigeria, FIN Finnish in Finland, GBR British in England and Scotland, GIH Gujarati Indian in Houston, GWD Gambian in Western Division, IBS Iberian populations in Spain, ITU Indian Telugu in the UK, JPT Japanese in Tokyo, KHV Kinh in Ho Chi Minh City, LWK Luhya in Webuye, MSL Mende in Sierra Leone, MXL Mexican Ancestry in Los Angeles, PEL Peruvian in Lima, PJL Punjabi in Lahore, PUR Puerto Rican in Puerto Rico, STU Sri Lankan Tamil in the UK, TSI Toscani in Italy, YRI Yoruba in Ibadan.

### GWAS of PTB and GA

We provided full GWAS results to allow access to non-European-ancestry summary statistics and facilitate future trans-ethnic GWAS meta-analyses of PTB and GA. As shown in Fig 2, we did not identify genetic associations with PTB or GA at genome-wide significance in our dataset. This result was somehow expected due to the appropriately QC’d but underpowered genetic dataset. When examining QQ plots, there was no indication of bias due to genomic inflation in the GWAS based on *λ* = 1.0 (Fig 2). We reported markers with suggestive association with PTB and GA at *P* < 1.0×10^-5^. In total, 8 SNPs were identified suggestively associated with PTB, most of them related with a lower risk of PTB (i.e., longer gestational age at delivery) per additional risk allele, except for two markers mapping to the *KDM4C* and *SOX5* loci. The strongest signal was observed in rs13151645 mapping to the non-coding RNA *LINC01182*. This SNP was associated with a 26% (95%CI= 0.65, 0.84) lower risk of PTB per additional risk allele, which was consistent with the association observed between rs13151645 and GA (/3= 0.6 weeks, 95%CI= 0.30, 0.91) (Table 2). Other SNPs in strong correlation (r^2^ > 0.6) with rs13151645 but with larger GWAS *P*-values, were seen in the nearby region (Fig 3), suggesting that *LINC01182* could be a good candidate locus for PTB study. Additionally, we observed a similar direction of association with GA at top signals detected in the GWAS of PTB, even though *P*-values were larger (i.e., less statistically significant).

**Fig 2.**
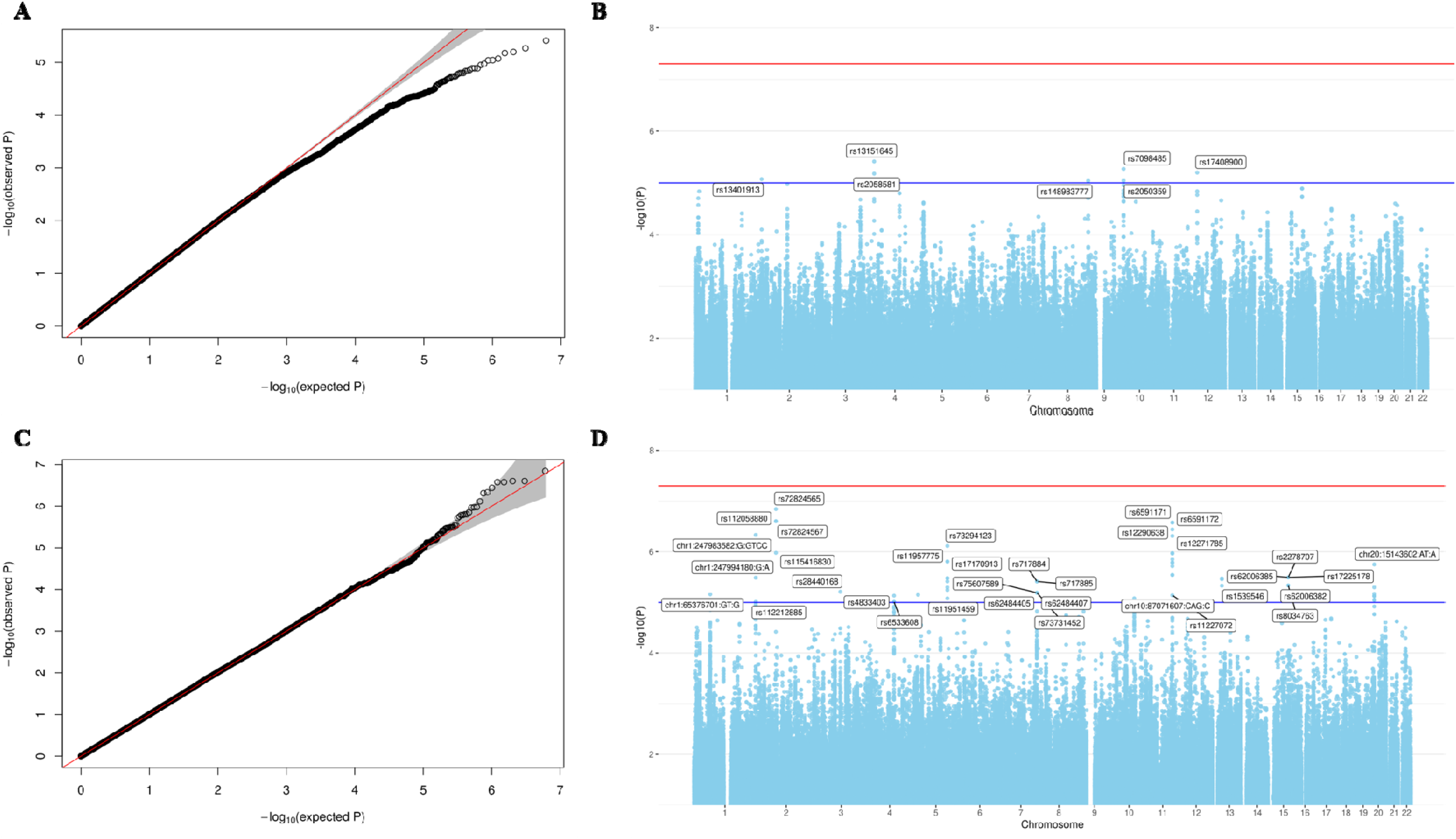
Quantile and Manhattan plots summarizing results of the GWAS of Preterm Birth (A, B) and Gestational age at delivery (C, D) conducted among women in the PAGE study (N=2,212). In the Manhattan plots (B, D), the horizontal blue line corresponds to the suggestive genome-wide association threshold at *P* < 1.0×10^-5^; the horizontal red line is the Bonferroni significant threshold at *P* < 5.0×10^-8^. Associations were adjusted for maternal age and the first two genetic PCs. Top SNPs were annotated with their rsID using the Haplotype Reference Consortium panel (GRCh37/hg19), or the SNP coordinates reported in TOPMED (GRCh38/hg38).

**Fig 3.**
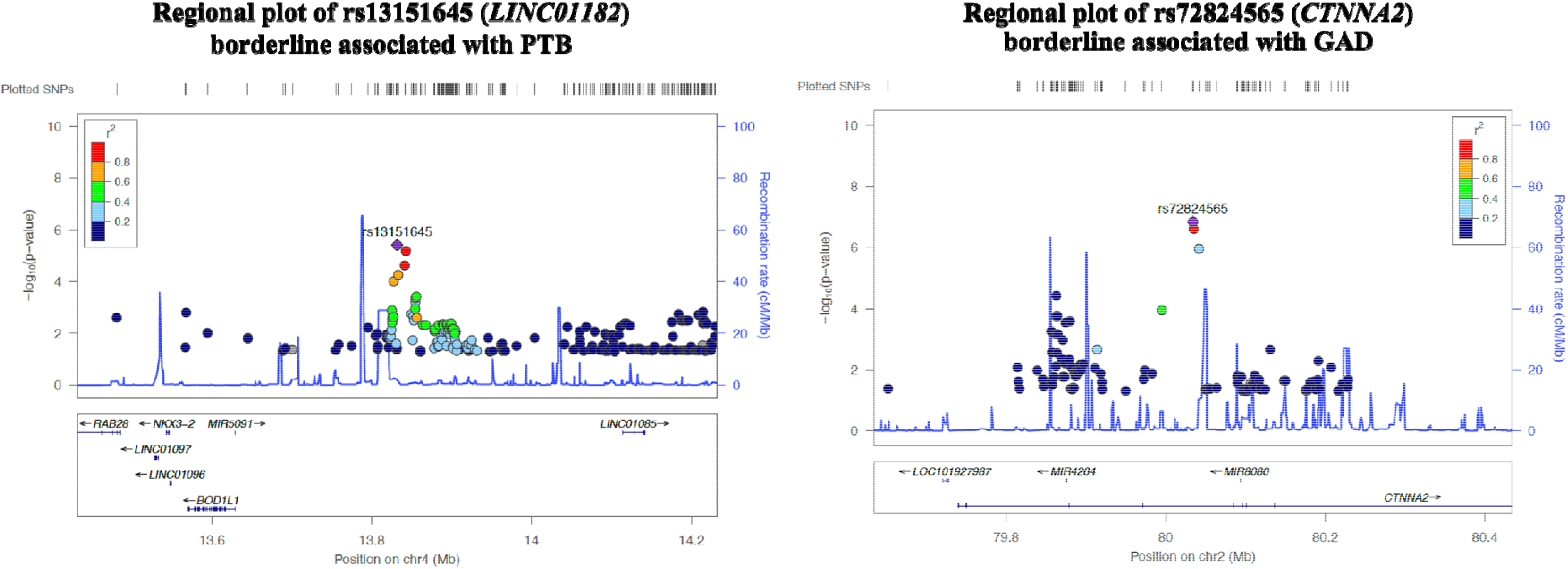
Regional plots showing the genetic context of top signals detected with suggestive association with Preterm Birth (rs13151645 in chr4) and Gestational age at delivery (rs72824565 in chr2). The colored legend represents the correlation (LD) o nearby SNPs with our sentinel SNP shown by the purple diamond. SNPs more correlated with the SNP of interest are shown as red dots. Pairwise SNP correlations were calculated using the 1,000 genomes (phase 3) and the admixed American population as reference. At the bottom of the plot, showing representative genes in the region and their transcriptional orientation.

**Table 2.** Top genetic associations detected in the GWAS of preterm birth (PTB) among women in the PAGE study (N=2,212). Suggestive associations were identified with *P* < 1.0×10^-5^.

|  |  |  |  |  | GWAS of PTB |  | Look up in GWAS of GA |  |
| --- | --- | --- | --- | --- | --- | --- | --- | --- |
| Gene and SNP | Ch<sub>r</sub> | Alleles (A1/A0) | Freq | $r^2$ | OR (95% CI) | P | $\beta$ (95% CI) | P |
| *MIR3681HG* |  |  |  |  |  |  |  |  |
| rs13401913 | 2 | T/C | 0.06 | --- | 0.52<br>(0.39,0.70) | $8.5 \times 10^{-6}$ | 1.32 (0.69,1.95) | $4.2 \times 10^{-5}$ |
| rs4849108 | 2 | G/A | 0.28 | $< 0.1$ | 0.74<br>(0.64,0.84) | $1.1 \times 10^{-5}$ | 0.56 (0.24,0.88) | $5.8 \times 10^{-4}$ |
| *LINC01182* |  |  |  |  |  |  |  |  |
| **rs13151645** | **4** | **T/T** | **0.65** | --- | 0.74<br>(0.65,0.84) | **$3.9 \times 10^{-6}$** | 0.60 (0.30,0.91) | $1.0 \times 10^{-4}$ |
| rs2058581 | 4 | T/T | 0.64 | 0.88 | 0.74<br>(0.65,0.85) | $6.6 \times 10^{-6}$ | 0.58 (0.28,0.88) | $1.8 \times 10^{-4}$ |
| *KDM4C* |  |  |  |  |  |  |  |  |
| rs148983777 | 9 | C/G | 0.07 | --- | 1.69<br>(1.34,2.13) | $9.1 \times 10^{-6}$ | -1.21 (-1.76,-0.65) | $1.9 \times 10^{-5}$ |
| *RP11-45P17.5: AKR1C3* |  |  |  |  |  |  |  |  |
| rs7098485 | 10 | G/G | 0.51 | --- | 0.76<br>(0.67,0.86) | $5.4 \times 10^{-6}$ | 0.56 (0.27,0.85) | $9.9 \times 10^{-5}$ |
| rs2050359 | 10 | A/A | 0.51 | 0.97 | 0.75<br>(0.67,0.85) | $9.1 \times 10^{-6}$ | 0.58 (0.29,0.87) | $1.8 \times 10^{-4}$ |
| *SOX5* |  |  |  |  |  |  |  |  |
| rs17408900 | 12 | C/T | 0.17 | --- | 1.44<br>(1.23,1.68) | $6.3 \times 10^{-6}$ | -0.70 (-1.08,-0.33) | $2.6 \times 10^{-4}$ |

In bold, SNP identified with the smallest P-value of association in the GWAS of PTB. Genome-wide significant signals if $P < 5.0 \times 10^{-8}$ . Associations were adjusted for maternal age (years) and the first two genetic PCs. SNP coordinates are based on the GRCh37hg19 genome build. A0: reference allele, A1: effect allele, Freq: frequency of the risk allele, $r^2$ : linkage disequilibrium (LD) showing the correlation between SNPs within the same genetic region. Pairwise LD correlations were calculated using the 1,000 genomes (phase 3) and the mixed American population as reference. Effect size is interpreted as the odds of PTB per additional risk allele. In the look-up of top signals of PTB in the GWAS of gestational age at delivery, the effect size is the estimated change in the number of gestational weeks, per additional risk alleles.

In the GWAS of GA (Table 3), 20 SNPs mapping to 12 genes across the genome were borderline associated (*P* < 1.0×10^-5^). For most of these markers, the risk allele investigated was associated with shorter GA at delivery, as it was evident for associations in the *DNAJC6*, *CTNNA2* and *GUCY1A2* loci. Positive associations with GA (i.e., longer gestational length) were identified in the *ALPK1:RTEL1P1*, *ARNT2,* and *MACROD2* genes. The marker most strongly associated with GA was detected in the intronic variant rs72824565 mapping within the *CTNNA2* gene (/3= -3.48 weeks, 95%CI= -4.78, -2.19). Few other signals in correlation with rs72824565 were seen in the region of *CTNNA2* (Fig 3). Top signals were nominally associated with shorter gestation, directionally consistent with their effect sizes in PTB GWAS.

**Table 3.**
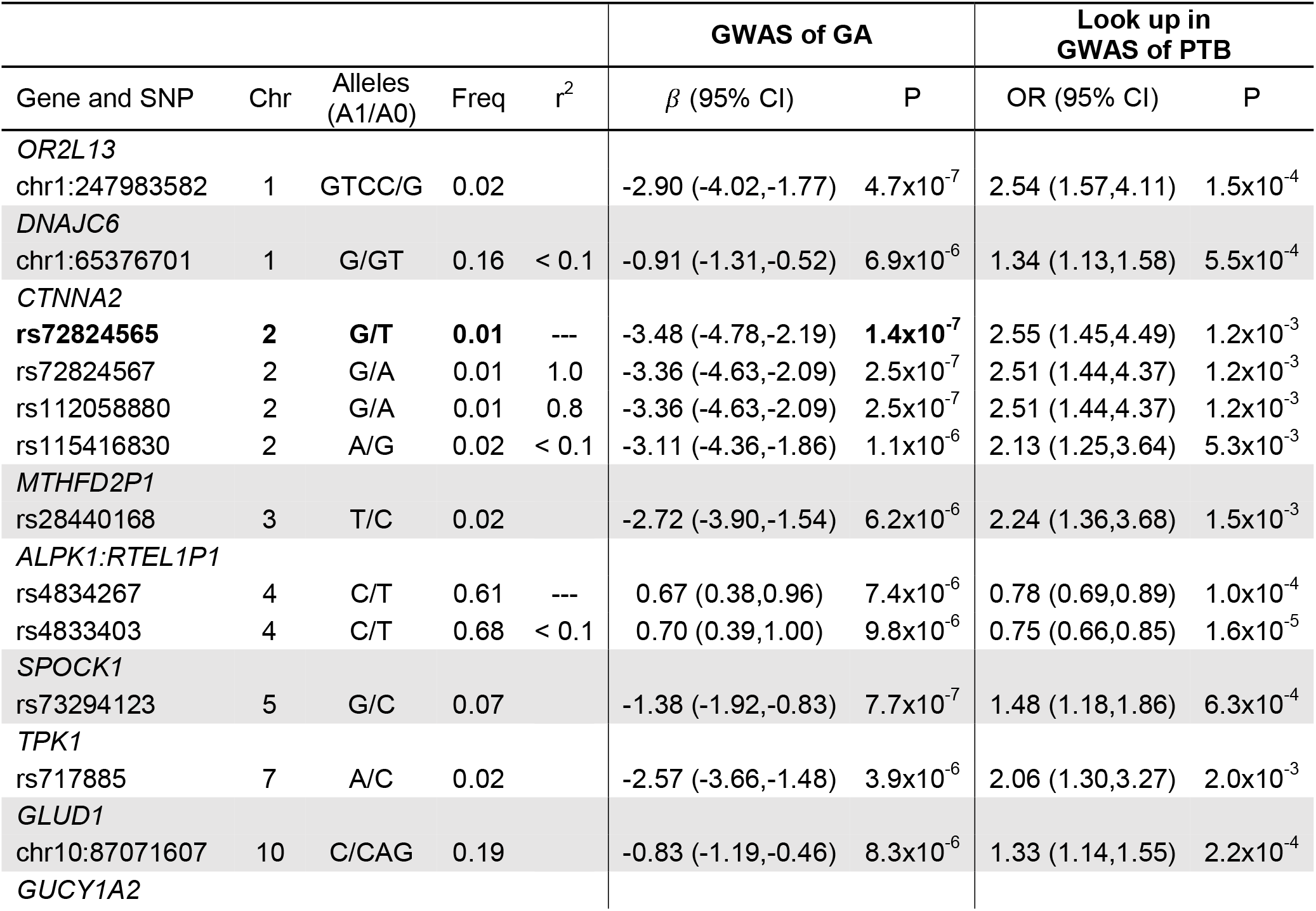
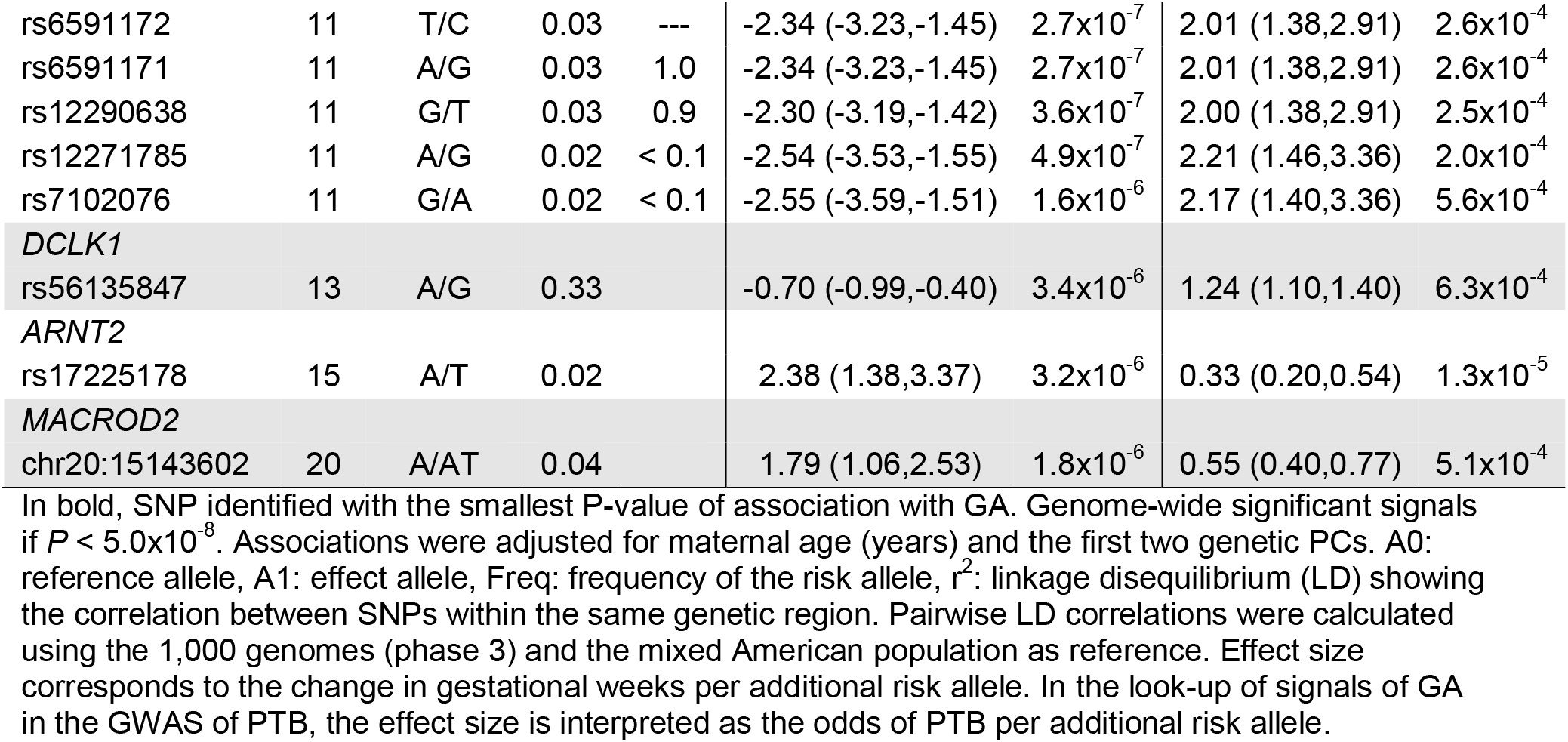
Top genetic associations detected in the GWAS of gestational age at delivery (GA, continuous) among women in the PAGE study (N=2,212). Suggestive associations were identified at *P* < 1.0×10^-5^.

Top independent signals identified in the GWAS in PAGE were looked up in a large meta-analysis of GWAS of PTB and GA conducted in Europeans [17]. We observed no replication of our signals in the GWAS in Europeans using a nominally significant GWAS *P* < 0.05. For top signals in PAGE identified in the GWAS in Europeans (7 and 16 top SNPs for PTB and GA, respectively), only a few of them (3/7 PTB and 1/16 GA SNPs) showed same direction of association between studies.

Using the Nagelkerke’s pseudo-R^2^ and the adjusted-R^2^ obtained from multivariable logistic and linear regressions, respectively, we assessed the percent of the total variance in PTB and GA explained by top SNPs detected with the smallest *P*-value in each GWAS. Combining the effect of the top 8 PTB-associated SNPs, they explained 7.2% of the variance in the risk of PTB in PAGE samples. This value was slightly larger than the variance explained by non-genetic risk factors (i.e., maternal age, gravidity, pre-pregnancy BMI, vaginal bleeding, maternal education, child sex, and two genetic PCs) (adjusted-R^2^= 6.9%) (S2 Table). Likewise, combining the effect of the top 20 GA-associated SNPs, they explained 10% of the GA variance, almost double the variance captured by non-genetic risk factors (adjusted-R^2^= 5.5%). Adding the effect of non-genetic and genetic risk factors, we captured 14% and 15.3% of the total variance in PTB and GA in our sample, respectively.

### Replication of European Markers and Genetic Score

We extracted summary statistics for 15 unique SNPs previously identified in association with PTB and/or GA in a meta-analysis of GWAS conducted among Europeans [16]. Out of the 11 GA SNPs reported by Zhang *et al.*, seven were observed in our dataset, but only three of them had the same direction of association with GA as in the reference study. True replication was only seen for two GA SNPs mapping to the *WNT4* gene (*P*=1.0×10^-3^) (Table 4). Similarly, out of the 6 SNPs previously reported in association with PTB, only two were identified in our dataset, both with the same direction of association with PTB as in the reference study, but with replication *P* > 0.03 (significant if *P* < 0.05/2 SNPs) (Table 4). We searched for proxies for SNPs in the reference study missing in our GWAS, but the two proxies (LD R^2^ > 0.7) in rs5950498 and rs5991030 identified for target SNPs rs5950506 (PTB) and rs5950491 (GAD) in Chr X, were also not included in our GWAS. Additionally, for each participant in PAGE, we constructed a standardized GRS for GA based on five independent SNPs previously identified by Zhang *et al*. (S3 Fig). We showed that an SD increase in the GRS was associated with an average 0.29- week (∼ 2 days) (95%CI= 0.08, 0.49 weeks) increase in GA (S3 Table). The GRS was able to capture 0.3% of the variance in GA in our sample, and in combination with other non-genetic risk factors (maternal age, parity, pre-pregnancy BMI, vaginal bleeding, maternal education, child sex, PC1, and PC2), it explained up to 6.0% of the variance in GA in our dataset.

**Table 4.** Look-up of risk loci for preterm birth and gestational duration identified in Europeans (Zhang *et al*. 2017), in the GWAS conducted in PAGE. SNP associations were replicated if a similar direction of association was observed between the reference and the observed GWAS SNP at a nominal GWAS *P-value* < 0.05.

| Reference GWAS in Europeans (Zhang et al. 2017) |  |  |  |  |  | GWAS in PAGE |  |  |
| --- | --- | --- | --- | --- | --- | --- | --- | --- |
| Gene | SNP | Alleles (A0/A1) | Chr: Position | $\beta$ | P | A1 | $\beta$ | P |
| *Association with GA* | | | | | | A1 | $\beta$ | P |
| *EBF1* | rs2963463 | C/T | 5:157895049 | -0.16 | $7.7 \times 10^{-24}$ | T | 0.01 | $9.6 \times 10^{-01}$ |
| | rs2946171 | T/G | 5:157921940 | -0.21 | $8.1 \times 10^{-21}$ | G | -0.02 | $9.3 \times 10^{-01}$ |
| *EEFSEC* | rs2955117 | G/A | 3:127881613 | 0.19 | $9.5 \times 10^{-15}$ | A | -0.04 | $8.1 \times 10^{-01}$ |
| | rs200745338 | D/I | 3:127869457 | 0.27 | $7.5 \times 10^{-16}$ | - | - | - |
| *AGTR2* | rs201226733 | I/D | 23:115164770 | -0.24 | $7.2 \times 10^{-16}$ | - | - | - |
| | rs5950491 | C/A | 23:115129714 | -0.25 | $6.6 \times 10^{-16}$ | - | - | - |
| ***WNT4*** | **rs56318008** | **C/T** | **1:22470407** | 0.32 | **$3.4 \times 10^{-14}$** | **T** | **0.48** | **$1.1 \times 10^{-03}$** |
|  | **rs12037376** | **G/A** | **1:22462111** | 0.34 | **$5.6 \times 10^{-14}$** | **A** | **0.48** | **$1.1 \times 10^{-03}$** |
| *ADCY5* | rs4383453 | G/A | 3:123068359 | -0.08 | $3.7 \times 10^{-08}$ | A | 0.20 | $3.6 \times 10^{-01}$ |
| | rs9861425 | A/C | 3:123072883 | -0.20 | $4.2 \times 10^{-10}$ | A | 0.20 | $2.4 \times 10^{-01}$ |
| *RAP2C* | rs200879388 | I/D | 23:13130057 | -0.16 | $3.4 \times 10^{-09}$ | | | |
| *Association with PTB* |  |  |  |  |  | A1 | OR | P |
| *EBF1* | rs2963463 | C/T | 5:157895049 | 1.13 | $4.5 \times 10^{-15}$ | T | 1.00 | $9.8 \times 10^{-01}$ |
| | rs2946169 | C/T | 5:157918959 | 1.16 | $2.2 \times 10^{-13}$ | T | 1.01 | $9.0 \times 10^{-01}$ |
| *EEFSEC* | rs201450565 | D/I | 3:128058610 | 0.82 | $1.9 \times 10^{-12}$ | - | - | - |
| | rs200745338 | D/I | 3:127869457 | 0.80 | $3.3 \times 10^{-14}$ | - | - | - |
| *AGTR2* | rs201386833 | D/I | 23:115164281 | 1.18 | $1.0 \times 10^{-11}$ | - | - | - |
| | rs5950506 | G/A | 23:115175748 | 1.18 | $1.1 \times 10^{-11}$ | - | - | - |

In bold, associations of SNPs in *WNT4* with GA that were replicated in the GWAS in PAGE at nominal *P* < 0.05. The effect estimate for GA is interpreted as the change in gestational weeks, per additional risk allele; positive values indicate longer gestational age at delivery associated with the SNP. For PTB, effect estimates are interpreted as the odds of PTB, per additional risk allele; values larger than 1 indicate increased odds of PTB associated with the SNP. SNPs in Zhang et al. without data in the GWAS in PAGE were filtered out during genetic QC.

### In-silico Functional Analysis

Based on the FUMA-GWAS analysis, five SNPs were identified as independent significant signals for PTB at *P* < 1.0×10^-5^; these signals were enriched in intronic and in non-coding RNAs within intronic regions (Fisher *P* < 0.01) (S1 Fig). Using the full distribution of GWAS *P*-values (*P* < 0.05), the MAGMA gene-set analysis identified a positive association of input PTB SNPs with GO terms associated with neuropeptide hormone activity, positive regulation of glucocorticoid secretion, inositol 1,3,4,5 tetrakisphosphate binding, and corticosterone secretion (adjusted-*P* < 2.0×10^-2^). Similarly, a tissue expression analysis identified a positive association between SNPs for PTB and genes expressed in adipose tissue, the heart, mammary breast cells, and the adrenal gland (*P* < 0.05). An eQTL look-up using BIOS QTL identified transcripts in *AKR1C3*, *AKR1CL1*, *AKR1C4*, among others, associated with the top PTB SNP rs7098485. Using prioritized genes (i.e., top 8 PTB SNPs at GWAS *P* < 1.0×10^-5^), differential gene expression in the above-mentioned tissues was identified for the *AKR1C3* gene associated with the SNP rs7098485. A gene-set enrichment analysis unveiled different biological processes related to the metabolism of polyketides, progesterone, glycoside, quinone, and steroids, in association with genes in *AKR1C3*, *AKR1C4*, *AKR1CL1*, all linked to the PTB SNP rs7098485.

*In-silico* interrogation of signals detected in the GWAS of GA revealed 15 independent significant SNPs (LD < 0.1, *P* < 1.0×10^-5^) and 229 GWAS-tagged SNPs in LD > 0.6 with the independent SNPs, mapping to 41 genes. These SNPs were enriched in intronic, intergenic, and non-coding RNAs within exonic and intronic regions (S2 Fig). Gene set analysis revealed a positive association of GA SNPs with GO terms related to nucleotide sugar transmembrane transporters activity and the positive regulation of catecholamines secretion. Two GA SNPs in rs4834267 and rs17225178 were identified as eQTLs of transcripts in the *ALPK1* and *ARNT2* genes, respectively, and data from the GWAScatalog revealed that rs4834267, rs73294123, and rs6591172, were previously reported in association with gut microbiota [23], hepatitis B [24], and genetic interactions in Alzheimer’s disease [25], respectively. Tissue-specific enrichment analysis showed that genes associated with GA SNPs were enriched for DEGs up-regulated in adipose tissue and the brain (S2 Fig), while gene-set enrichment analysis revealed enrichment of GA SNPs for cancer modules and cancer gene neighborhoods (S4 Table).

## DISCUSSION

This may be the largest study investigating the maternal genetic factors associated with PTB and GA in South American samples. Our study included data from the PAGE study which were 2,212 women, 933 of whom were PTB cases. None of the ∼ 6 million SNPs interrogated surpassed significance in our analysis; however, the top signals identified with *P* < 1.0×10^-5^ were associated with a lower risk of PTB (n = 6/8 top SNPs), and with shorter GA (n = 16/20 top SNPs) in the specific GWAS analyses. Two SNPs in *WNT4* associated with GA in Europeans in a previous study were replicated in our sample (*P* < 0.001), and a GRS for GA constructed using five European SNPs, was associated with a 2-day longer GA in our study. The GRS explained 0.3% of the variance in GA, and this value was lower than the variance explained by the top 20 SNPs detected in our GWAS of GA (adjusted-R^2^= 10%), suggesting that PTB genetic risk factors from European-ancestry populations may not be generalizable to non-European, admixed American populations. Functional annotation of top markers identified in the GWAS provided insights into their possible biological role in PTB risk and GA duration based on maternal genetic effects.

Our strongest signal for the GWAS of PTB (rs13151645), maps to the long intergenic non-coding RNA (lincRNAs) 1182 (*LINC01182*), which was suggestively associated with a 26% lower risk of PTB and explained 1.2% of the variance in PTB in our sample. No previous evidence, to our knowledge, supports the role of rs13151645 or the *LINC01182* gene in PTB risk. However, several SNPs in the *LINC01182* gene in our data had a strong correlation (LD > 0.5) with our index SNP rs13151645, suggesting that this region may be of biological importance for PTB. For instance, Zhou *et al*. [26] identified five lincRNAs associated with circadian rhythm genes or “clock genes” that were downregulated in the placentas of women with spontaneous PTB versus controls. Similarly, these lincRNAs were linked to pathways relevant to PTB (immunity, inflammation, oxidative stress, apoptosis, etc.) [26]. Replication of our top signal at *LINC01182* in a larger dataset is necessary to confirm its potential mechanistic role in PTB risk.

Another variant of interest observed with a nominal association was rs7098485 near the Aldo-Keto Reductase Family 1 Member C3 (*AKR1C3*). Other SNPs in high LD with rs7098485 were observed in this locus (n = 11) in our data. *AKR1C3* is a protein-coding gene member of the superfamily of aldo/keto reductases, which are enzymes that catalyze the conversion of aldehydes and ketones to their specific alcohols using NADH/NADPH as cofactors [27]. This enzyme is involved in the metabolism of sex hormones (estrogen, androgens, and progesterone) and prostaglandins [28], which are acidic lipids that promote inflammatory responses [29]. Prostaglandins are ubiquitously produced by different mammalian cells [29, 30], and during gestation, they are known to promote labor by stimulating smooth muscle contractility in the uterus and cervical ripening [2, 14, 30, 31]. Thus, the role of *AKR1C3* on PTB may be through alterations in the function of this enzyme that result in a reduction in the biochemical availability of prostaglandins. This is consistent with our observation of *AKR1C3* SNPs having effect sizes suggestive of protective effects on PTB. This concept may be additionally supported by the previous identification of an eQTL associated with our SNP rs7098485, where the T risk allele (same risk allele for PTB) was associated with lower expression of the transcript ENSG00000196139 in *AKR1C3* (FDR < 0.001). To confirm our hypothesis of a potential functional role of rs7098485 on *AKR1C3* in PTB risk, a formal functional analysis is required. The expression of *AKR1C3* has also been associated with better survival in patients with endometrial cancer [32, 33], and with other diseases like breast cancer [28] and asthma [34].

Our *in-silico* functional analysis using top PTB-associated SNPs supported the role of these markers in the metabolism of progesterone, glycoside, and prostanoid (i.e., prostaglandin), primarily attributed to *AKR1C3* function. Another relevant pathway for PTB identified in our gene set analysis was linked to the regulation of steroid secretion (glucocorticoids and corticosterone); these are compounds commonly used as an antenatal treatment to improve pregnancy outcomes in women at risk of preterm delivery [35]. Our risk factor analysis demonstrated that the top 8 SNPs explained a slightly larger proportion of the variance in PTB compared to non-genetic risk factors (7.2% vs 6.9%), indicating the potential of genetic markers in adding value to the risk prediction of PTB already achieved by common risk factors. Of note, results of this genetic variance analysis should be interpreted with caution, as variance in GA and PTB was calculated in the same sample used to conduct the GWAS; thus, results may be overestimated. Furthermore, the SNPs included in the calculation of the phenotypic variance were only suggestively associated with the traits in our GWAS. None of the markers previously detected for PTB in Europeans were replicated in our sample (*P* > 0.03), and calculating a GRS was not possible as only one out of six SNPs previously reported in Europeans (rs2963463 in *EBF1*), surpassed QC for the GRS in our dataset.

The intronic SNP rs72824565 mapping to the Catenin Alpha 2 (*CTNNA2*) gene was the most significant signal in the GWAS of GA. This variant was associated with a 3-week shorter gestational duration, and this association was consistent across the three other GWAS SNPs observed in high correlation with rs72824565 in *CTNNA2*. The influence of this protein-coding gene on lower GA may be through fetal neurodevelopmental impairments. Specifically, *CTNNA2* acts as a link between the cadherin adhesion receptors and the cytoskeleton in regulating cell-cell adhesion and differentiation in the nervous system [36]. It also participates in cortical neuronal migration and neurite growth [36]. Bi-allelic loss-of-function mutations in *CTNNA2* have been associated with pachygyria syndrome, a neurodevelopmental brain defect associated with impairments in neuronal migration, resulting in lower gyri complexity in the cerebral cortex [36]. This syndrome is accompanied by motor and cognitive delays in affected patients [36]. Our findings at *CTNNA2* are consistent with the pathophysiology of PTB, as infants born preterm are more likely to suffer from neurodevelopmental disorders and long-term cognitive and motor disabilities [2, 14]. Other variants in *CTNNA2* have been previously associated with educational attainment [37], externalizing behaviors (attention deficit hyperactivity disorder, substance abuse, antisocial behavior) [38], and acute myeloid leukemia [39], among others. Another SNP detected with suggestive inverse association with GA in our study was located in chr1:65376701, mapping to the *DNAJC6* gene, a member of the heat shock family of proteins (HSP). SNPs mapping to this gene, and similar members of this family of proteins, have been vinculated with higher risk of sPTB [40]. This multi-omic study conducted in Europeans identified SNPs and transcripts of HSPs associated with higher sPTB, hypothesizing that activation of these proteins mediated signalling may promote labor [40], either through their role in activating immune reponses as a consequence of infection, or through their role in maturation and activation of nuclear hormone receptors, like glucocorticoid, androgen, estrogen and progesterone receptors [40].

For additional top SNPs identified in the GWAS of GA, the *in-silico* functional analysis suggested enrichment of their mapping genes for cancer modules in relation to SNPs in *ARNT2*, *DCLK1,* and *SPOCK1*. Furthermore, top GA-SNPs were associated with the positive regulation of catecholamines secretion in the gene-set analysis (*CHRNB2, GDNF, CXCL12, KCNB1*). Catecholamines are stress hormones that have been previously associated with an increased risk of spontaneous PTB [41]. GA-associated SNPs also showed upregulation of differentially expressed genes in adipose tissue, brain caudate, and brain putamen regions in the basal ganglia, supporting the concept that GA variants may be associated with PTB risk through their influence on fetal neurodevelopmental traits. Our risk factor analysis suggested that top SNPs associated with GA explained almost double the variance in the trait captured by common non-genetic risk factors (10% vs 5.5%), and this variance was close to the one reported by Zhang *et al.* in their GWAS (10% for us vs 17% in their study).

We replicated two associations in *WNT4* previously identified in Europeans, and the GRS using five European SNPs was associated with a 2-day increase in GA in our sample; the GRS captured 0.3% of the variance in GA in PAGE participants, which was less than the variance explained by our top SNPs of the GWAS. The replication of markers in *WNT4* across populations is worth noting, as it indicates that this locus may represent a common mechanistic pathway for PTB, which causal protective effect on the disease still needs to be validated. Biologically, *WNT4* is related to the decidualization of the endometrium for subsequent implantation and establishment of pregnancy [16].

Our study had several limitations. First, we did not differentiate PTB cases into two clinical presentations as PPROM or spontaneous PTB, limiting our ability to identify markers more targeted to the pathophysiological mechanisms of each subtype of PTB. The main reason why we did not do this stratified analysis was due to sample size constraints, as having smaller comparison groups would have been disadvantageous for the GWAS knowing the very large sample sizes required for this type of analysis (∼100k). In addition, we did not consider in our analysis the fetal genetic effects, knowing that GA may be influenced by both, maternal and fetal genetic effects [2]. However, previous studies looking at both genotypes coincided with the major influence that the maternal genotype has on GA and PTB determination versus the fetal genotype [16]. Lastly, our study may have been limited by the population studied, and by its specific socioeconomic and environmental context, with findings that may not be generalizable to other populations, as has been demonstrated in other trans-ethnic GWAS of PTB [15].

Despite these limitations, our study has several strengths. First, this is the largest GWAS of PTB and GA conducted among South American individuals, which expands the literature on the genetics of pregnancy outcomes in low-and-middle income populations from non-European ancestry. By making our GWAS results available, we facilitate their inclusion in future meta-analyses of GWAS that are more tailored to South American and other Latino and Hispanic populations; thus, promoting the discovery of genetic-based treatments that are more useful to these minority populations. As another strength, we used the most up-to-date reference panel for the genetic imputation to include the largest number of SNPs to be interrogated in the GWAS. Even though our results were null, we conducted different post hoc analyses to evaluate the biological meaning of the top signals identified in each GWAS. Lastly, we performed a replication analysis of signals previously identified in Europeans, and we developed a GRS for GA in PAGE samples based on European markers.

Further validation of our findings in larger datasets of similar ancestral backgrounds, may help in elucidating the mechanisms associated with the pathophysiology of PTB in the context of minority and vulnerable populations, and to identify genetic markers that serve in the early prediction of PTB in women at higher risk.

## CONCLUSIONS

Our study identified genetic markers suggestively associated with PTB and GA among Peruvian women. These markers were independent of those previously reported in Europeans, and they were linked to pathways related to the metabolism of steroids, progesterone, and prostanoids (*AKR1C3*), fetal neurodevelopment (*CTNNA2*), response to stressors (*DNAJC6*), and catecholamine secretion (*CXCL12*). Prioritized genes for GA were upregulated in metabolically relevant tissues such as adipose tissue and the brain, supporting some of our findings in the pathway analysis. Replication of suggestive markers in larger studies of the same population background is warranted to validate their role in PTB and GA. The discovery of new markers for PTB and the validation of existing ones will aid in developing genetic scores that permit an accurate risk stratification of women, especially in high-risk non-European ancestry populations.

## METHODS

### Study Population and Analytic Sample

This study was conducted among participants of the Placental Abruption Genetic Epidemiology study (PAGE), a case-control study from Lima, Peru, designed to investigate the genetic and environmental determinants of placenta abruption and other adverse pregnancy outcomes like preterm birth. Study procedures have been described elsewhere [42, 43]. Briefly, women were recruited from seven participating hospitals in Lima between March 2013 and December 2015. Preterm women were identified by daily monitoring of admission logbooks from antepartum wards, the emergency room (intensive care units), delivery wards, and surgery rooms [42].

Eligible women for this study were older than 18 years of age, had singleton pregnancies, had sufficient phenotypic and medical information to determine PTB status, and provided a saliva sample at delivery to extract genomic data (described below). Women were excluded if preterm deliveries were medically indicated. The total number of participants remained 933 PTB cases and 1,279 controls. Eligible women were invited to participate in a 30-minute in-person interview during their hospital stay, where research staff explained the study objectives and obtained written informed consent from study participants. All the study protocols were approved by the IRB of participating hospitals, and the Swedish Medical Center, Seattle, WA, where the study was administratively based.

### Data collection

Trained research staff conducted in-person interviews to gather information on the participants’ sociodemographic, lifestyle and anthropometric characteristics using structured questionnaires. Sociodemographic factors collected included maternal age, education level (higher vs lower than high school), marital status (married living with a partner vs not married living alone), employment status, health perception during pregnancy (good to excellent vs fair to poor), and infant sex. Lifestyle characteristics included smoking or alcohol use during pregnancy, and drug abuse. Pre-pregnancy body mass index (BMI) was measured continuous and categorically (underweight if BMI < 18 kg/m^2^, normal weight if BMI 18-24.9 kg/m^2^, overweight if BMI 25-30 kg/m^2^, and obese if BMI 30-49 kg/m^2^). Maternal obstetric variables were abstracted from medical records and included gestational age at delivery (weeks), mode of delivery (natural vs C-section), planned pregnancy, diagnosis of preterm premature rupture of fetal membranes (PPROM), gravidity (primiparous vs multiparous), preeclampsia, vaginal bleeding, spontaneous abortions, and infant birthweight (g).

### Gestational Age at Delivery and Preterm Birth

Gestational age at delivery (GA) was obtained from the maternal self-reported date of the last menstrual period (LMP) during the interview. When values were missing for LMP, information on GA was obtained from an early pregnancy ultrasound or fundal height examinations performed ≤ 20 weeks of gestation according to medical records. We defined preterm births (PTB) following the American College of Obstetrics and Gynecologists criteria as deliveries occurring < 37 weeks of gestation [44]. PTB cases can be further classified into three pathophysiological groups: spontaneous PTB (sPTB), preterm with premature rupture of fetal membranes (PPROM), or medically induced PTB [45]. Women with sPTB have a medical diagnosis of spontaneous labor with intact fetal membranes prior to completing 37 weeks of gestation.

Women with PPROM have a physician diagnosis of premature rupture of fetal membranes before labor and prior to completing 37 weeks of gestation. Of the 933 PTB cases in the study, 527 (56%) were sPTB and 379 (41%) were PPROM. A high proportion of C-sections was seen in both clinical subgroups (70% in sPTB and 68% in PPROM). Controls were selected at random from women delivering at or after 37 weeks and before 42 weeks across the participating hospitals. By study protocol, controls were captured within 48 hours of the identification of a PTB case. To preserve a larger smple size for genetic analyses and increase power [46], we run the GWAS using the general PTB definition and including both clinical subgroups.

### DNA Extraction and Genotyping

At delivery, maternal saliva samples were collected, plated, and stored at room temperature using the Oragene^TM^ saliva cells kit (OGR500, DNA Genotek, Ottawa, Canada) [47, 48]. Genomic DNA was extracted using the Qiagen DNAeasy^TM^ system and following manufacturer protocols (Qiagen, Valencia, CA) [47]. Direct genotyping of ∼ 300,000 genome-wide genetic variants was performed at the Genomics Shared Resource, Roswell Park Cancer Institute, Buffalo, NY, using the Illumina HumanCore-24 BeadChip array (Illumina Inc., San Diego, CA) [47, 48].

### Data Quality Control and Imputation

Prior to performing genetic imputation, we conducted different quality control (QC) steps on the directly genotyped genetic variants or SNPs. Firstly, we filtered out SNPs based on minor allele frequency (MAF) < 0.01 (n=39,439), missing genotyping rate > 5% (n=19,700), and deviation from the Hardy-Weinberg equilibrium (HWE *P* < 1.0×10^-5^, n=223) in the control group. After quality control, 249,299 directly genotyped SNPs remained in the dataset. Similarly, samples were filtered out if they had genotyping failure rate > 0.1 (n=38), were duplicates or related considering an identity by descent (IBD) value > 0.9 (n=20), had excess heterozygosity/homozygosity rate (outside the range of ± 3 standard deviations from the mean heterozygosity; n=29), and had discordance between the reported sex and sex predicted using genetic information (F ≥ 0.8; n=10). We did not exclude one sample identified with ancestry divergence (outside the following range for the first two genetic principal components (PCs): PC1 > -0.01 and < 0.01, and PC2 > -0.02 and < 0.02). This sample was retained knowing the ethnical homogeneity of our dataset (most are mestizos - or mixed race/ethnicity), and because we included the first two genetic PCs as covariates to account for potential population stratification in downstream analyses (see below). A total of 2,212 samples (933 cases and 1,279 controls) were retained for further analyses. We then performed genetic imputation to infer the genotype of SNPs not included in the array and to maximize our chances of identifying true genetic associations with PTB and/or GA. First, we used the Michigan server (https://imputationserver.sph.umich.edu) [49] to conduct a pre-phasing step and check for inconsistencies between our input genotype, and data from the 1000 Genomes phase 3 (version 5) Admixed American (AMR) population used as the reference panel. Cleaned pre-phased genotypes were then phased using EAGLE (version 2.4) [50] to infer haplotypes and improve imputation accuracy [47]. Imputation of phased genotypes was conducted using the TOPMed imputation server (https://imputation.biodatacatalyst.nhlbi.nih.gov/) and reference panel (version r2). Imputed genotype data was then QCed to exclude SNPs that were non-biallelic SNPs, duplicated, had quality imputation score (INFO) < 0.8, HWE *P* < 1.0×10^-5^, MAF < 0.01, and genotyping call rate < 99%, leaving in total 6,047,004 high-quality SNPs for the analysis. SNPs were annotated to their rsID (dbSNP142, GRCh37/hg19) using data from the Haplotype Reference Consortium (HRC) reference panel (HRC.r1-1) [51], or using the chromosome and position if the rsID was not available.

### Ancestry Analysis

Using directly genotyped SNPs that passed quality control, we performed a combined genetic PC analysis between samples from PAGE and the 1000 Genomes Project (phase 3, version 2013.05.02) to determine the genetic ancestry of participants. First, we generated PCs for SNPs identified in common between the 1000 Genomes and PAGE datasets. Then, we retrieved genetic variation captured by the first three PCs obtained from the combined dataset analysis and used scatterplots to visually represent the clustering of samples based on their genetic relatedness across pairs of PCs (i.e., PC1 vs PC2 and PC1 vs PC3). We also generated PCs for the PAGE dataset alone and retrieved the first two PCs, explaining in total 66.1% of the total genetic variation in the sample, to adjust for population stratification in subsequent analyses.

### Statistical Analysis

We described characteristics of study participants using the mean and standard deviations (SD) for continuous variables, and proportions and percentages for categorical variables.

Comparisons between groups (PTB cases and controls) were done using t-tests (Wilcoxon rank sum test if non-parametric) and chi-squared tests for continuous and categorical variables, respectively. Using an additive genetic model (i.e., assuming a linear increase in PTB risk/GA per additional risk allele), we fitted multivariable logistic and linear regression models to test the association of each SNP as the exposure against PTB and GA as the outcome, respectively, using PLINK (version 1.90) [52]. Genome-wide associations at each SNP were adjusted for maternal age, and the first two genetic PCs to account for population stratification. We used the genomic inflation factor or *λ* to identify population stratification or residual confounding in the GWAS if *λ* > 1.0. Adjusted regression coefficients (ORs/Betas), 95% confidence intervals, and the genomic control uncorrected *P*-values (∼ to genomic controlled corrected *P*-values) were reported for each SNP. To determine if there was consistency in the genetic effect between traits, we looked up association estimates for top signals (*P* < 1.0×10^-5^) identified in the GWAS of PTB, in the GWAS of GA, and vice versa. Similarly, we looked up our top suggestive signals in the largest and most recent European GWAS meta-analysis conducted to date for GA and PTB [17] (http://egg-consortium.org/Gestational-duration-2023.html). We applied correction for multiple testing (6,047,004 tests performed) using the Bonferroni method and regarded GWAS significant associations at *P* < 5.0×10^-8^, or nominally significant associations at *P* < 1.0×10^-5^.

Results of the GWAS were summarized using Manhattan and Quantile plots (Q-Q plots). We used stepwise adjusted regressions to obtain the percent of the total variance in the traits explained by each one of the top SNPs (*P* < 1.0×10^-5^) identified in the GWAS of PTB and GA, and by all of them in combination. We reported the adjusted-R^2^ obtained from the linear regressions between the SNPs (exposures) and GA (outcome), or the Nagelkerke’s pseudo-R^2^ obtained from the logistic regressions between the SNPs and PTB. Similarly, we estimated the percent of the total variance in the traits (GA and PTB) explained by the non-genetic risk factors of maternal age, gravidity, pre-pregnancy BMI, presence of vaginal bleeding, maternal education, child sex, and the first two genetic PCs. We then compared the proportion of the variance in the traits explained by the genetic, the non-genetic risk factors, and by the combination of both factors.

### Replication of European Markers and Polygenic Score

We retrieved summary statistics for genome-wide significant associations obtained in a recent meta-analysis of GWAS of PTB and GA conducted by Zhang *et al.* in European cohorts (N=52,211, spontaneous PTB cases with GA < 37 weeks = 5,896 and term controls = 46,315) [16]. Across the discovery and replication stages of this metanalyses, 11 SNPs were identified in association with PTB (in 6 genes), 6 with GA (in 3 genes), and 2 SNPs were identified in common between both traits [16]. We looked up associations for these 15 unique SNPs in the GWAS conducted in PAGE and considered evidence of replication if a similar direction of association was identified between studies with replication *P* < 0.05/number of SNPs interrogated. Since effect estimates were expressed in days for SNPs associated with GA in Zhang *et al.*, we divided them by 7 to match the interpretation of GA used in our study (in weeks). For SNPs in Zhang *et al.* [16] not present in our GWAS, we searched for proxy SNPs with an LD R^2^ > 0.7 with the target SNP using LDlink (https://ldlink.nih.gov/) [53] and the 1000 genomes as the reference panel (AMR and EUR populations). Using individual-level genotype data in PAGE and summary statistics from Zhang *et al.*, we developed a genetic risk score (GRS) for each participant in PAGE using PLINK (version 1.90). The score for each participant represents the sum of the total number of risk alleles present in their genotype, weighted by the effect of each SNP in the score as obtained from GWAS summary data. SNPs from the reference study were clumped to include only independent SNPs (LD < 0.1) in the score, resulting in five independent GA-associated SNPs but only one independent PTB-associated SNP; thus, we only constructed the GRS for GA using PAGE samples. The score was standardized to represent a weekly change in GA per SD increase in the GRS. The variance (R^2^) in GA explained by the standardized score was obtained using the adjusted-R^2^ from a full model (with the score), minus the adjusted-R^2^ from a basic model including only covariates (maternal age, gestational age, PC1, and PC2).

### In-silico Functional Analysis

We used summary data from GWAS conducted among PAGE participants to perform an *in- silico* functional analysis using tools available in the FUMA-GWAS (Functional Mapping and Annotation of GWAS) web tool (https://fuma.ctglab.nl/) [54]. This analysis was restricted to associations with *P* < 0.05 in the GWAS. For gene annotation purposes, we specified as the reference panel the 1000 Genomes AMR population, selected an LD threshold < 0.6 to identify independent SNPs, and chose a *P* < 1.0×10^-5^ as the minimum *P*-value to detect lead SNPs.

Other settings were left as default. FUMA-GWAS uses two applications, the SNP2GENE (version 1.3.5d) and the GENE2FUNC (version 1.3.5), to provide complete SNP annotation and biological interpretation of GWAS data, respectively. Using the SNP2GENE function, we reported information on the lead SNP in a region, the number of independent SNPs (r^2^ < 0.1) and their functional consequences (intronic, exonic, non-coding RNA, etc.), the nearest gene, and output from the MAGMA analysis. For this latter, gene set and tissue-specificity analyses were performed using curated data obtained from the MsigDB (version 7.0) and GTEx (v7) databases, respectively. When relevant, we also reported phenotypic data obtained from the GWAScatalog and expression Quantitative Trait Loci (eQTL) retrieved from the BIOS QTL [55], regarding lead SNPs in our dataset. Using genes prioritized in the SNP2GENE analysis, we run the GENE2FUNC function to test the enrichment of candidate genes for curated gene sets obtained from MsigDB, WikiPathways, and reported genes in the GWAScatalog. Enrichment was also tested for tissue-specific gene expression using data from GTEx (v8) based on pre- calculated differentially expressed gene sets (DEGs). In both cases, enrichment analysis was performed using hypergeometric tests, with correction for multiple testing using the Bonferroni method. Further information on the protocol implemented in the FUMA-GWAS analysis can be found on their website (https://fuma.ctglab.nl/tutorial).

## ACKNOWLEDGEMENTS

The authors are grateful to the participants of the collaborating hospitals for their involvement in the study. We also want to recognize the important contribution of Ms. Elena Sanchez and the research staff of the Asociacion Civil Proyectos en Salud (PROESA), Peru, for their expert technical assistance with this project.

## AUTHOR CONTRIBUTIONS

DLJQ and BG contributed to the conception and design of the study. LL and TW provided their expert feedback on the methodology of the study and the interpretation of results. DLJQ conducted the analyses and wrote the paper in collaboration with BG. SES contributed with access to the data. SES, LL, TW, and MAW provided their expert clinical, epidemiological, and statistical input on the manuscript. All authors read the paper and agreed with the version submitted for publication.

## DATA AVAILABILITY

Complete summary data of the GWAS of GA and PTB would be available from the GWAS catalog EMBL-EBI repository. Study accession numbers will be released upon approval of the manuscript for publication.

## SUPPORTING INFORMATION

**S1 Table. Pregnancy history and maternal mental health characteristics of study participants (N=2,212).**

**S2 Table. Variance in preterm birth explained by top SNP with the smallest P-value detected in the GWAS in PAGE, and by known non-genetic risk factors.**

**S3 Table. Variance in gestational duration (GAD) explained by top SNP with the smallest P-value detected in the GWAS in PAGE, and by known non-genetic risk factors.**

**S4 Table. Enrichment analysis showing pathways associated with genes annotated to top SNPs detected in the GWAS of preterm birth and gestational duration in PAGE.**

**S1 Fig. Functional analysis of top-five independent (linkage disequilibrium < 0.1) genetic variants nominally associated with preterm birth (PTB) at *P* < 1.0×10^-5^.**

**S2 Fig. Functional analysis of top-12 independent (linkage disequilibrium < 0.1) genetic variants nominally associated with gestational duration (GAD) at *P* < 1.0×10^-5^.**

**S3 Fig. Distribution of the polygenic risk score for gestational duration among study participants (N=2,212).**

